# Implementation of Virtual Reality Pain Alleviation Therapeutic into Routine Pediatric Clinical Care: Experience and Perspectives of Stakeholders

**DOI:** 10.1101/2024.03.19.24304228

**Authors:** Helen Girin, Megan Armstrong, Kim A. Bjorklund, Christopher Murphy, Julie B. Samora, Jonathan Chang, Daniel J. Scherzer, Henry Xiang

**Affiliations:** Center for Pediatric Trauma Research, The Abigail Wexner Research Institute, Nationwide Children’s Hospital, 700 Children’s Drive, Columbus, OH 43205, USA; Center for Injury Research & Policy, The Abigail Wexner Research Institute, Nationwide Children’s Hospital, 700 Children’s Drive, Columbus, OH 43205, USA; Section of Plastic Surgery, Nationwide Children’s Hospital, 700 Children’s Drive, Columbus, OH 43205, USA; Department of Plastic Surgery, The Ohio State University College of Medicine, 370 West 9^th^ Avenue, Columbus, OH 43210, USA; Department of Orthopedics, Nationwide Children’s Hospital, 700 Children’s Drive, Columbus, OH 43205, USA; Division of Emergency Medicine, Nationwide Children’s Hospital, 700 Children’s Drive, Columbus, OH 43205, USA; Department of Pediatrics, The Ohio State University College of Medicine, 370 West 9^th^ Avenue, Columbus, OH 43210, USA

**Keywords:** Pediatric, Virtual Reality, Acute Pain, Anxiety, Implementation

## Abstract

**Aims:** To determine the feasibility of implementing virtual reality (VR) in three pediatric clinical environments during brief, painful procedures outside of research.

**Design:** A quality improvement project with quantitative and qualitative feedback between March and November 2023.

**Methods:** Medical providers (doctors and nurses) implemented VR during brief pediatric medical procedures and completed a demographics and feasibility survey. Qualitative data were obtained from semi-structured interviews after the implementation period.

**Results:** Patients (n=30) played the VR game during either their medically necessary pin-pulling or needlestick procedures within three clinical environments. Children ranged from 5-16 years and were 50% male. The majority of patients reported enjoyment (mean 8.2 out of 10) with the VR during the procedure and only one minor technical issue was reported. Qualitative data collection showed the benefits of using VR included its ease of use, decreased observed anxiety, and patients had an easier time getting through the medical procedures.

**Conclusion:** Clinician feedback from the dissemination of VR into pediatric clinical environments showed promising results. Standardized guidelines are needed to further implement VR pain alleviation as standard care in clinical settings.

**Implications for patient care:** VR is easy to implement in clinic settings and can improve pediatric patient care during painful medical procedures. Utilizing nurses as champions for novel clinical techniques can assist with transitioning from research to the standard of care.

**Impact:** The project provided evidence for broader expansion and implementation of VR into different clinical areas. If VR is implemented into daily clinical practice, patients could benefit from reduced pain and anxiety, and medical procedures could be performed more easily than without adjunctive pain/anxiety management.

**Reporting Method:** This project adhered to the Standards for Reporting Qualitative Research (SRQR) checklist.

**Patient or Public Contribution:** No Patient or Public Contribution

**What does this paper contribute to the wider global clinical community?:**

- Medical providers (including nurses) were able to effectively implement VR for pain and anxiety distraction without extending procedure time.
- Nurses are excellent champions for implementing novel techniques for patient care.

**Trial and Protocol Registration:** There is no trial and protocol registration for this project. This project evaluated the feasibility of medical providers implementing VR outside of research. Thus, a structured protocol or trial was outside the scope of the project.

**Statistics Statement:** The authors have checked to make sure that our submission conforms as applicable to the Journal’s statistical guidelines. There is a statistician on the author team (Dr. Henry Xiang). The authors affirm that the methods used in the data analyses are suitably applied to their data within their study design and context, and the statistical findings have been implemented and interpreted correctly. The authors agree to take responsibility for ensuring that the choice of statistical approach is appropriate and is conducted and interpreted correctly as a condition to submit to the Journal.

## 1. Introduction

Pain is an important component of clinical care and requires thoughtful pain management strategies to avoid psychological and emotional strain for patients and family members. Acute pain is “the physiologic response and experience to noxious stimuli that can become pathologic, is normally sudden in onset, time limited, and motivates behaviors to avoid actual or potential tissue injuries.”^1^ Opioid pain medications are commonly prescribed to treat acute pain following injury or medical procedures, however, there has been a push to decrease the amount of opioids prescribed due to the United States opioid crisis, including state cap laws.^3^ The United States Department of Health and Human Services named the opioid epidemic in 2017 as a public health emergency, and has renewed this declaration yearly thereafter.^2^ Notably, in the pediatric population, it has been shown that opioid exposure post-surgery is associated with persistent opioid use among opioid-naïve patients.^4^ Non-pharmacological pain management methods are particularly relevant as they can address acute pain needs while sparing opioid side effects and the potential for misuse.

Music, movies and games have been used in US pediatric clinical settings to distract from painful procedures.^5^ Virtual Reality (VR) has been shown to be an effective pain distraction tool during pediatric clinical care including needlestick procedures^6,7^ and burn dressing changes.^8,9^ Our team developed a virtual reality pain alleviation therapeutic (VR-PAT) to address the procedural pain experienced during pediatric burn dressing changes in the outpatient clinic.^8^ This VR-PAT was a fun, child-appropriate VR game developed at our institution with feedback from patients and clinicians and was hosted on an iPhone and displayed on a low-cost VR headset. Considerations were made to ensure the system was lightweight and highly portable to ensure it was practical for a clinical setting. In our previous randomized controlled trial, participants using the active VR-PAT self-reported clinically meaningful lower pain scores than those who only received the standard of care.^8^ Nurses also reported that the VR-PAT was easy to implement in the clinic and found it to be helpful for the majority of dressing changes.^8^ Through further analysis, we found a link between anxiety and pain, with self-reported anxiety prior to the dressing change being significantly associated with self-reported pain following the procedure.^10^ Soumil et al. further discovered that the perceived realism, fun, and engagement of VR-PAT significantly impact the effectiveness of these interventions for reducing pain.^11^ Finally, our team found a clinically significant reduction in self-reported pain while using the VR-PAT during home care of burn injuries.^12^ None of our participants reported serious adverse events, which supported our hypothesis that VR is a safe non-pharmacological pain alleviation therapeutic.

While VR has been found safe and effective for acute pain alleviation during pediatric procedures, there are gaps in the dissemination and clinical implementation of these novel interventions outside of research domains. Researchers provided evidence that it can take decades to move evidence of effectiveness in the research domain to clinical implementation.^13^ Through research dissemination efforts, our research team was approached by clinicians to trial our VR-PAT in their clinical settings. The purpose of this project was to report our experience having medical staff in the Emergency Department (ED), Plastic Surgery Clinic, and Orthopedic Clinic implement VR-PAT.

## 2. Methods

### Study design and population

Multiple clinical providers at a large academic pediatric hospital approached the research team about the feasibility and implementation of using VR-PAT in their clinical settings (Figure 1). The research team had separate meetings with each of the medical providers that would be involved in this study to discuss our VR-PAT research, their goals, and expectations of the VR-PAT. The Institutional Review Board (IRB) at Nationwide Children’s Hospital reviewed this project and determined that the proposed activity was not research involving human subjects as defined by Department of Health and Human Services and Food and Drug Administration regulations, and waived approval. The requirement of consent/assent was waived by the IRB. Following the IRB review, an in-person meeting was scheduled with each medical provider to demo the headset and answer any questions. A survey was provided to gather some brief de-identified information about how the procedure went while using the VR-PAT. The research team did not have any clinical involvement with the implementation of the VR other than assisting with any questions or problems should they arise. The research team also collected the surveys from each of the medical providers. Completed surveys were scanned and sent to a point person on the research team after each participant encounter or at the end of the month if prompted. Survey data were entered into a Research Electronic Data Capture (REDCap)^14,15^ database. After each clinic completed a convenience sample of 10 surveys (n=30 total), feedback meetings were scheduled to collect qualitative data about the experience.

**Figure 1.**
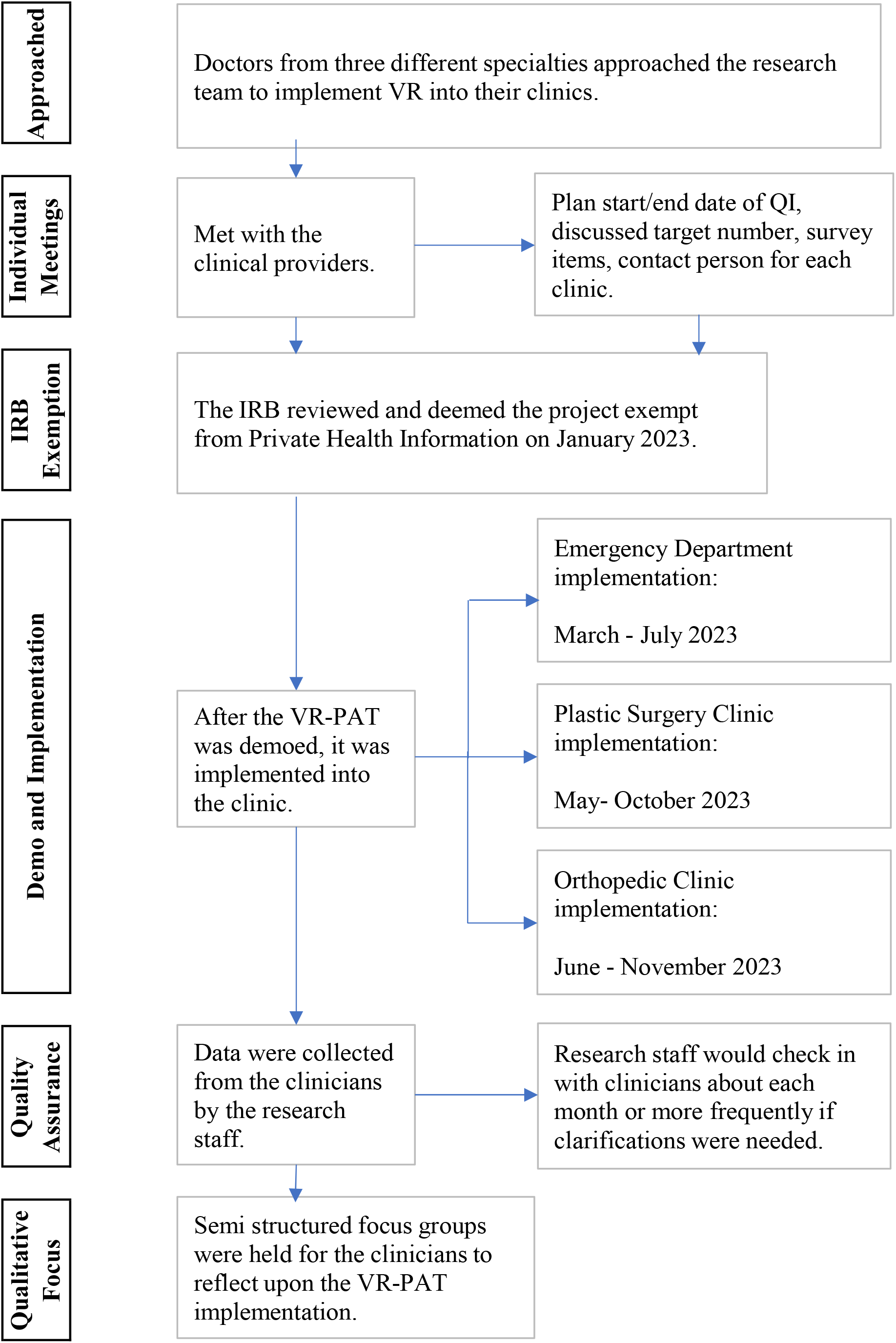
Flow chart of VR-PAT implementation and evaluation.

Eligible Participants were children between the ages of 5 and 17 years of age who spoke English and did not have a medical history of seizures or adverse reactions to VR.

### VR game

Our published research used the smartphone version of the VR-PAT app; however, we have since transferred this app onto a head-mounted display (PICO Neo3 Pro Eye). Each clinic was provided one VR PICO headset with the Virtual River Cruise game already downloaded. The game gave the child a first-person view of floating gently down a river in a boat with a penguin companion. The child moved their eyes from side to side to see a winter landscape and crystals that would appear within the field of vision. Looking directly at the crystal allowed it to be broken open to receive points on the scoreboard and a lower temperature on the thermostat. The difficulty of breaking the crystals increased with the length of time spent playing the game. The Virtual River Cruise game was played by having children slightly tilt their heads (no hand, finger, or arm involvement), which minimized interference with the medical procedure. The enjoyability and immersive environment were designed to create a calming virtual environment to reduce anxiety. The game automatically started when the headset was powered on to limit additional steps for medical staff and lasted indefinitely so that the VR-PAT could be played throughout the entire medical procedure with a fully charged device. Details of the smartphone-based VR-PAT have been described in previous publications.^8,11^

### Survey Questions

The medical care providers filled out a survey for each participant (supplementary file 1). Survey items included: age of patient, sex, month and year of VR-PAT use, patient reported VR-PAT enjoyment, and technical issues with using the VR.

Patient-reported VR-PAT enjoyment was obtained by asking “On a scale of 0-10, how much did you like playing the VR-PAT game during your procedure?” (0=did not like it at all, 10=liked it the most possible). Upon completion of the medical procedure, the medical provider reported any technical issues with using the VR-PAT.

### Post Implementation Focus Group

After each clinic collected a total of 10 surveys, a semi-structured interview was scheduled with the implementing clinician and the research team. These individual interviews were meant to collect qualitative feedback about the clinic’s experience implementing the VR-PAT with their patients. The following questions were shared to capture the common themes: “Describe the experience of using the VR at your clinic”, “What do you think about the VR that worked well in the clinic setting?”, “What do you think about the VR did not work well in the clinic setting?”, “Do you have any suggestions for improvement, whether it be the survey or the VR game?”, “In your opinion, what should we do to disseminate into other clinics or hospitals?”

## 3. Results

### VR-PAT clinical Implementation

Males made up 50 % of the study population with a mean age of 9.8 years (range 5-16 years) (Table 1). Children from the ED were younger than children in the Orthopedic & Plastic Surgery clinics who needed pin-pulling (mean 7.8 and 10.9 years, respectively). Children across the study reported enjoying the VR-PAT (mean 8.2, range 0-10). The pin-pulling cohort reported one technical problem where the patient had problems popping the crystals. This was a minor issue that was resolved by restarting the headset.

**Table 1.** Clinical Implementation of VR-PAT for Quality Improvement.

|  | **Total**<br>**N=30** | **ED**<br>**N=10** | **Pin-Pulling**<br>**N=20** |
| --- | --- | --- | --- |
| Age, Mean (range) | 9.8 (5-16) | 7.8 (5-14) | 10.9 (6-16) |
| Male, n (%) | 15 (50%) | 6 (60%) | 9 (45%) |
| Female, n (%) | 15 (50%) | 4 (40%) | 11 (55%) |
| Enjoyment Score*, Mean (range) | 8.2 (0-10) | 7.5 (0-10) | 8.7 (3-10) |
| Technical Issues, n (%) | 1 (3%) | 0 (0%) | 1 (5%) |

\*How much did you like playing the VR game during your procedure? (0=did not like at all, 10=liked it the most possible)

### Qualitative Outcomes

Table 2 reports common themes from the semi-structured interviews with medical providers.

**Table 2:** Benefits and Challenges of Using VR-PAT from Semi-Structured Interviews.

| ***Benefits*** | ***Challenges*** |
| --- | --- |
| Patient had an easier time with medical procedure | There are children who are too anxious to accept VR-PAT |
| Medical providers perceived less anxiety in patients | Needs more gamification/competition for older kids |
| Easy to use after explanation to medical staff | VR-PAT was designed for six-year-olds and above |
| Easy to introduce and explain the game to the patients |  |
| Little time required to set up the headset/game |  |
| Integrating it into the clinic flow was not difficult |  |
| Can be played for the entirety of the medical procedure |  |
| Minor technical difficulties could be resolved by medical staff |  |

All three clinics commented that they saw signs of children being less anxious. They reflected upon children’s anxiety without VR-PAT with patients pulling away from the procedure and parent reassurance. Medical providers noted that when the child was fully immersed in the VR-PAT, they would remain still, and the parent would usually have minimal interaction with the child. However, some kids were too anxious to allow visual obstruction and needed to see what the medical staff were doing during the procedures. These patients were not able to be fully immersed into the VR environment.

Features of the VR-PAT allowed the medical providers to use it in their clinical setting without prolonging the procedure time. By having the Virtual River Cruise game start immediately when the VR headset was powered on and the game playing continuously allowed the child to be fully immersed into the game during the whole procedure. The game was also easy to introduce and explain to patients, so they did not have a steep learning curve. There was little technical assistance required from the research team after having a scheduled demonstration on how to use and implement the VR-PAT.

A challenge medical providers encountered was that game seemed to work better for younger children, as older children expressed the desire for a more challenging or competitive game. Also, since the game was designed and tested in children aged 5-17 years, this limited the ability to recruit patient’s younger than five years old.

## 4. Discussion

This project reports very promising results of implementing VR-PAT into clinical settings outside of a research domain. While this project has a small convenience sample, we were able to provide evidence that this intervention was easy to implement without impeding the required medical procedure. Children also reported enjoyment playing the game, with clinicians noting an observed decrease in anxiety before and during the procedures.

Participants in this project were evenly distributed between sexes (50% female) and covered a range of ages (5-16 years). Only one technical issue was reported that was easy to resolve within the clinic without outside assistance. This is important for further clinical implementation of VR-PAT. Orthopedic clinics report continued use of VR-PAT six months after data collection ended. Most children reported that they enjoyed playing the game (8.2 out of a maximum 10). This enjoyment score ranged from 0 to 10, which is not unusual. A few patients in our prior VR research also expressed a desire to watch the procedure and did not like having their eyes covered. Additionally, the Virtual River Cruise game was designed for a broad age range and to support many skill levels. Because of this, some older patients and those who have more video game experience could feel that the game is too easy and needs more gamification to create a more challenging experience. This clinical implementation is reflective of prior VR-PAT research in which most children report enjoying playing VR during medical procedures.^8,12^

During the semi-structured interviews, clinicians notably mentioned that patients had an easier time with the medical procedures. Clinicians talked about how without VR, patients were very anxious prior to the pin-pulling or needlestick procedures and the anxiety often added time to the procedure with parents and medical staff offering encouragement. We did not include a metric to capture anxiety in our survey, but anecdotally, clinicians reported that when patients were fully immersed in the VR-PAT early in the appointment, they were able to complete the procedure more quickly and without additional reassurance. Clinicians found it difficult to comment on observed pain, as these pin-pulling and needlestick procedures are very short but felt that the decreased anxiety may have helped with perceived pain as well. Other researchers have found similar results during pediatric medical procedures. Kilic et al, found a statistically significant reduction in anxiety after a burn dressing among patients using the VR intervention as compared to the control group and more satisfaction with their medical care.^16^ Gold et al, also found significantly lower patient and clinician reported anxiety scores post peripheral intravenous catheter placement in the VR group as compared to a standard of care group.^17^ The qualitative evidence provided in the semi-structured interviews warrants more formal research into VR-PAT’s impact on procedural anxiety.

This project should be interpreted within the context of several limitations. First, this clinical implementation was made up of a small convenience sample from three clinics at one pediatric hospital. This sample limits the generalizability of findings, however, since the project purpose was to determine the feasibility of implementing VR-PAT in real-world clinical settings using a hand-off strategy, this population was appropriate for our study goals. Second, a limitation of full implementation was that the VR-PAT was designed for children age 5 years or older, so younger children were excluded. It can be difficult to immerse young children into a developmentally appropriate VR and who can have trouble fully communicating their emotions. Our team recognizes the high anxiety needs of younger children and are exploring innovative distraction methods for future research. Third, some medical providers reported that they are prone to motion sickness and could not wear the VR-PAT and experience the game for themselves.

Therefore, they had to rely on researcher’s explanation of the game to guide the patient through the game. This is important feedback for further implementation considerations, but since we had success with this project, we do not feel that this impacted our results. Finally, we tried implementing our VR-PAT into different clinical settings, without the same success. This was an early implementation prior to some important VR-PAT changes. We learned some lessons from this early implementation and made important VR-PAT changes prior to this implementation. These changes included the game starting as soon as the headset was turned on and finding a single contact person in the clinic to be a project champion.

## 5. Conclusion

In conclusion, the dissemination of VR-PAT in real-world pediatric clinics and the feedback provided by the clinicians implementing the VR-PAT showed positive and very promising results. The research and clinical domains need to work together to create standardized guidelines for VR-PAT to be implemented as standard of care in clinical settings. Standardized VR guidelines will offer a stronger argument for insurance reimbursement for medical VR usage.

## Data Availability

All data produced in the present study are available upon reasonable request to the authors

## Acknowledgments

This study was supported the Agency for Healthcare Research and Quality (R01 HS29183-01, PI-Henry Xiang). The funder had no involvement in study design; in the collection, analysis and interpretation of data; in the writing of the report; and in the decision to submit the article for publication.

