## Supplementary File 1 for "Implementation of Virtual Reality Pain Alleviation Therapeutic into Routine Pediatric Clinical Care: Experience and Perspectives of Stakeholders"

### Clinic Survey

| | |
|---|---|
| Record ID | ________________________________ |
| Clinical Location | ◯ Burn Clinic<br>◯ ED<br>◯ Orthopedic (pin-pulling) |
| Age of Patient | ________________________________<br>(not birthdate) |
| Sex | ◯ Male<br>◯ Female<br>◯ Undisclosed |
| Month of VR use | ◯ January<br>◯ February<br>◯ March<br>◯ April<br>◯ May<br>◯ June<br>◯ July<br>◯ August<br>◯ September<br>◯ October<br>◯ November<br>◯ December |
| Year of VR use | ◯ 2022<br>◯ 2023<br>◯ 2024 |
| On a scale of 0-10, How much did you like playing the VR game during your procedure? (0=did not like at all, 10=liked it the most possible) | ________________________________<br>(for patient to answer) |
| Were there any technical issues with using the VR? | ________________________________________<br>(for nurse to answer) |

REDCap®
